# Health Insurance and Maternal Healthcare Utilisation in Nepal: Evidence from the Nepal Demographic and Health Survey 2022

**DOI:** 10.64898/2026.09.29.26364291

**Authors:** Yash Raj Lamsal, Abdulaziz Mohammed Hussen, Joyce L. Browne, Reshu Agrawal Sagtani

## Abstract

**Background:** Access to maternal healthcare services is essential for improving maternal and neonatal health outcomes. Health insurance can reduce financial barriers to healthcare access and may facilitate utilisation of maternal health services, and contextualized evidence can further support policy development and implementation. This study assessed the association between health insurance coverage and utilisation of antenatal care (ANC), institutional delivery, skilled birth attendance and postnatal care (PNC) services among women in Nepal.

**Methods:** This study used data from the 2022 Nepal Demographic and Health Survey (NDHS), a nationally representative cross-sectional survey. The analysis included women aged 15–49 years who had a live birth within the 24 months preceding the survey. Inverse probability of treatment weighting (IPTW) based on propensity scores was applied to balance measured socioeconomic and demographic characteristics between insured and uninsured women. Weighted marginal structural models were used to estimate the association between health insurance coverage and maternal health-care utilisation outcomes. Robustness was assessed using alternative propensity score specifications and E-values for unmeasured confounding.

**Results:** Among 14,845 women interviewed, 2,078 reported a recent live birth and were included in the analysis (unweighted; 1,998 when survey-weighted). Of these, 201 (9.7%) had health insurance coverage and 1,877 (90.3%) were uninsured. After adjustment using IPTW, health insurance coverage was associated with significantly higher utilisation of maternal healthcare services. Insured women had more than twice the odds of attending at least four ANC visits compared with uninsured women (AOR 2.14; 95% CI 1.15–3.98). Similarly, insured women had higher odds of receiving a PNC check-up within two days of delivery (AOR 2.05; 95% CI 1.28–3.27) and receiving PNC from a skilled provider (AOR 1.94; 95% CI 1.23–3.06). Although insurance coverage was positively associated with institutional delivery (AOR 1.55; 95% CI 0.91–2.62) and skilled birth attendance (AOR 1.57; 95% CI 0.90–2.72), these associations did not reach statistical significance.

**Conclusion:** Health insurance coverage was associated with greater utilisation of antenatal and postnatal healthcare services among women in Nepal. However, no significant association was observed for institutional delivery or skilled birth attendance. Expanding health insurance coverage, particularly among underserved populations, may help improve maternal healthcare utilisation and support progress towards universal health coverage and maternal health-related Sustainable Development Goals.

## 1 Introduction

Nepal has made substantial progress in reducing maternal mortality over the past two decades, largely due to improvements in maternal healthcare services and increased coverage of essential interventions across the continuum of care (Thapa, 2021; Exemplars in Global Health, 2020). Maternal mortality declined considerably up to 2015; however, significant challenges remain in achieving the Sustainable Development Goal (SDG) target of fewer than 70 maternal deaths per 100,000 live births by 2030. The Nepal Maternal Mortality Study, conducted alongside the 2021 National Population and Housing Census, estimated the maternal mortality ratio at 151 deaths per 100,000 live births (Ministry of Health and Population (Nepal) and National Statistics Office (Nepal), 2022). More recent modelled estimates from the UN Maternal Mortality Estimation Inter-Agency Group place Nepal’s maternal mortality ratio at 142 deaths per 100,000 live births in 2023 (World Health Organization et al., 2025). The two figures are not directly comparable: the former is a direct estimate from a census-based mortality study, the latter a modelled estimate derived from multiple data sources. Leading causes of maternal mortality include haemorrhage, hypertensive disorders of pregnancy, sepsis, and complications arising from unsafe abortions. These risks are further compounded by sociocultural factors such as early marriage, delayed care seeking, and limited access to skilled birth attendants, particularly in rural and geographically remote areas (Sitaula et al., 2021; Mahara et al., 2016).

To improve maternal and newborn health outcomes, the Government of Nepal has integrated SDG targets into its periodic development plans, most recently the Sixteenth Plan (2024/25–2028/29), which continues the priority given to maternal, neonatal, and child health services (National Planning Commission (Nepal), 2024). Key strategies include expanding healthcare infrastructure, strengthening the health workforce, improving access to skilled birth attendance and emergency obstetric care, and increasing coverage of antenatal care (ANC) and postnatal care (PNC) services, particularly in underserved areas. Evidence from Nepal indicates that increased utilisation of ANC, skilled birth attendance, PNC, and emergency obstetric care has contributed to reductions in maternal mortality (Adhikari et al., 2022; Pandey et al., 2025; Adhikari et al., 2025).

Financial barriers remain an important determinant of maternal healthcare utilisation. Evidence from low- and middle-income countries (LMICs) suggests that health insurance is associated with increased utilisation of maternal health services (Kazibwe et al., 2024; Wang et al., 2017; Anindya et al., 2020). To improve financial protection and progress towards universal health coverage, Nepal introduced the National Health Insurance Program (NHIP), administered by the Health Insurance Board (HIB). The scheme functions as a social health protection mechanism in which the HIB acts as the purchaser of services and health facilities operating under the Ministry of Health and Population provide care (Khanal et al., 2023). NHIP covers a broad range of health services, including maternal, newborn, and child health services such as ANC, skilled delivery care, PNC, child immunisation, and routine health check-ups. By reducing out-of-pocket expenditure, the programme aims to improve equitable access to essential healthcare services.

Despite these objectives, enrolment in health insurance remains low in Nepal and dropout rates are high (Khanal et al., 2023). Previous studies have identified several contributing factors, including limited awareness of insurance benefits, perceived inadequacies in service quality, administrative barriers, and dissatisfaction with the benefit package (Ranabhat et al., 2020; Acharya et al., 2023; Khanal et al., 2023). According to the 2022 Nepal Demographic and Health Survey (NDHS), 88% of women and 87% of men reported having no health insurance coverage. Among insured individuals, most were enrolled in the government insurance programme, although private insurance schemes also operate in Nepal and provide coverage to a smaller proportion of the population (Ministry of Health and Population (Nepal) et al., 2023). Given the low overall coverage and potential inequities in enrolment, understanding whether health insurance is associated with improved maternal healthcare utilisation is important for informing future policy and programme design.

Yet, evidence from Nepal remains limited and has not fully explored recent patterns of service utilisation using the latest nationally representative data. Therefore, this study assesses the association between maternal health insurance coverage and the utilisation of maternal healthcare services, specifically ANC, institutional delivery, skilled birth attendance, and PNC, using data from the 2022 NDHS. The findings will inform strategies to improve access to maternal healthcare services and support Nepal’s progress towards achieving its maternal health and universal health coverage goals.

## 2 Methods

### 2.1 Study design

This study used data from the 2022 NDHS, a nationally representative cross-sectional household survey. The manuscript was prepared in accordance with the Strengthening the Reporting of Observational Studies in Epidemiology (STROBE) guidelines for observational studies (von Elm et al., 2007), with details in Supplementary Table S6. The NDHS collects information on demographic characteristics, fertility, family planning, maternal and child health, nutrition, and healthcare utilisation across Nepal. The survey employed a two-stage stratified cluster sampling design to ensure national representativeness across all seven provinces and ecological regions.

### 2.2 Study settings

The NDHS 2022 was conducted throughout Nepal, including the provinces of Koshi, Madhesh, Bagmati, Gandaki, Lumbini, Karnali, and Sudurpashchim, covering Mountain, Hill, and Terai ecological regions. Details of the survey methodology have been published elsewhere (Ministry of Health and Population (Nepal) et al., 2023).

### 2.3 Participants

Women aged 15–49 years who reported a live birth within the 24 months preceding the survey were eligible for inclusion. Restricting the analysis to recent births minimised recall bias and ensured that information on ANC and PNC utilisation reflected recent healthcare practices and health insurance coverage. Where women reported multiple births during the reference period, only the most recent birth was included to maintain independence of observations. The NDHS interviewed 14,845 women aged 15–49 years. After restricting the sample to women who had experienced a recent live birth within the previous 24 months, the final analytical sample comprised 2,078 women. A participant flow diagram is presented in the Results section (Figure 2).

### 2.4 Study variables

#### Outcome variables

Maternal healthcare utilisation was assessed using three primary outcome indicators: attendance of at least four ANC visits during the most recent pregnancy, receipt of a PNC check-up within two days of delivery, and receipt of PNC from a skilled healthcare provider. Institutional delivery and skilled birth attendance were examined as secondary outcomes in order to describe utilisation across the full continuum of care. These are reported alongside the primary outcomes in Table 2 and are labelled as secondary throughout. All outcome variables were coded as binary measures (yes/no). The threshold of four ANC visits was selected because it corresponds to Nepal’s operational national standard. Under the Aama Programme, a cash incentive is paid to women on completion of four ANC visits at four, six, eight and nine months of pregnancy, so ANC4+ is the indicator against which national maternal health performance is monitored and financially incentivised (Ministry of Health and Population (Nepal), 2019). Although the World Health Organization recommends at least eight antenatal contacts (World Health Organization, 2016), Nepal has not yet rolled out this model. The Safe Motherhood and Newborn Health Road Map 2030 says eight visits are too ambitious for now. It directs that the focus stay on four good-quality visits, while working towards eight over the Road Map period (Ministry of Health and Population (Nepal), 2019). Four visits therefore remains the national benchmark and the standard measure in DHS-based analyses.

#### Exposure variables

The primary exposure variable was health insurance coverage. Women were classified as insured if they reported enrolment in either a government or private health insurance scheme at the time of the survey. Because the number of women covered by private insurance schemes was relatively small within the NDHS sample, government and private insurance were combined into a single binary indicator of insurance coverage. Health insurance was hypothesised to improve utilisation of maternal healthcare services by reducing financial barriers to accessing care.

#### Covariates

Potential confounders were identified through a review of the literature and informed by a directed acyclic graph (DAG). The DAG was specified using DAGitty (Textor et al., 2011) and rendered in R using ggplot2. DAGs provide a systematic framework for mapping causal relationships and identifying variables that should be adjusted to reduce bias (Greenland et al., 1999; Hernán and Robins, 2020). Based on prior studies of maternal healthcare utilisation in Nepal and other LMICs (Anindya et al., 2020; Were et al., 2017), the candidate confounders represented in the DAG were maternal age, birth order, educational attainment, household wealth status, marital status, occupation, place of residence (urban or rural), province of residence, decision-making autonomy, recent internet use, and perceived difficulty in accessing healthcare due to distance (Figure 1). Variables considered to be on the causal pathway between health insurance coverage and maternal healthcare utilisation were excluded to minimise the risk of over-adjustment bias (Pearl, 2009; Greenland and Robins, 2009).

**Figure 1:**
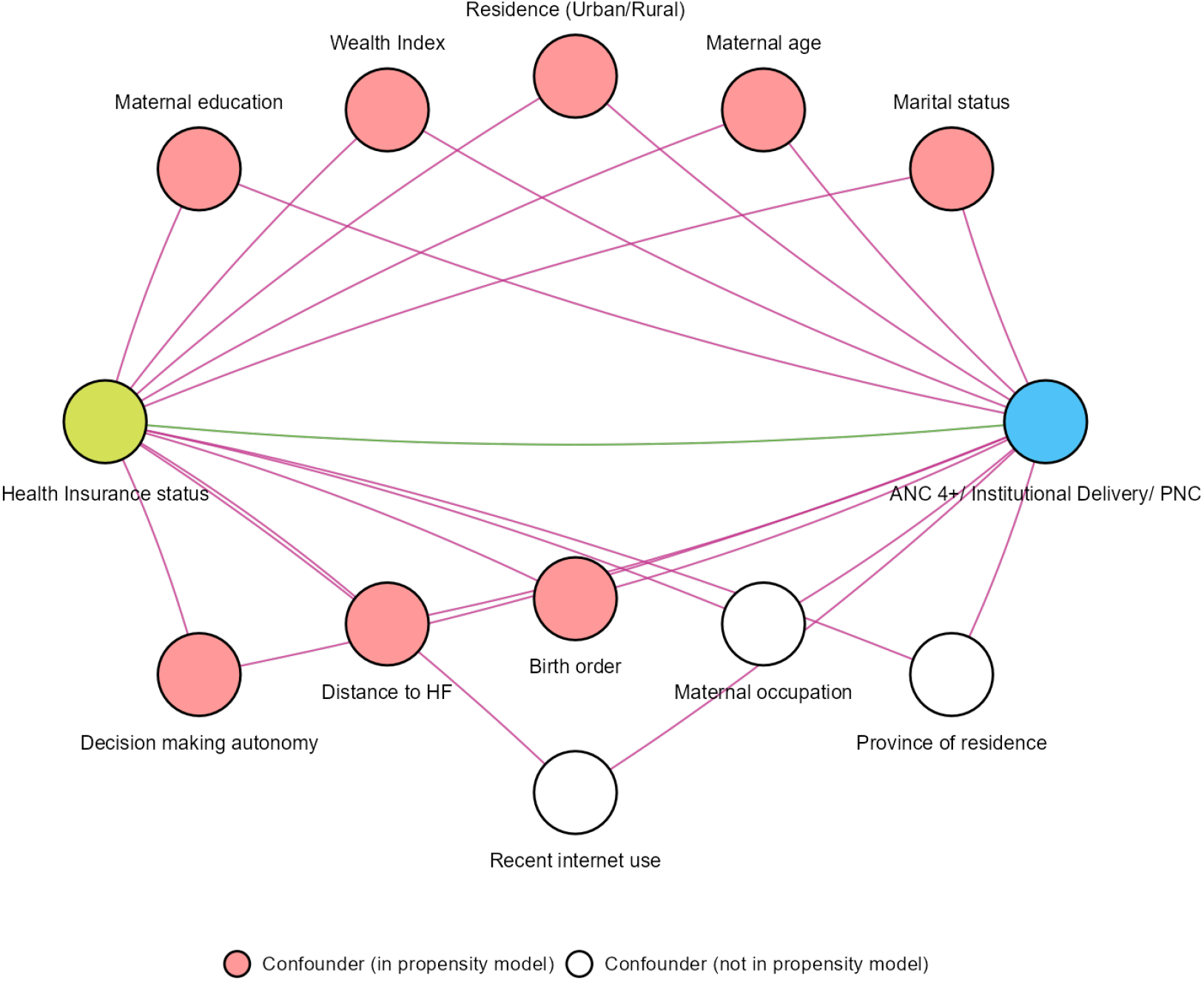
Directed acyclic graph for mapping relevant study variables. Shaded nodes are the eight confounders entered into the propensity score model; unshaded nodes (maternal occupation, province of residence, and recent internet use) were identified as candidate confounders but were not fitted, for the reasons given in the text. Supplementary Table S3 reports the estimates obtained when each is added back.

**Figure 2:**
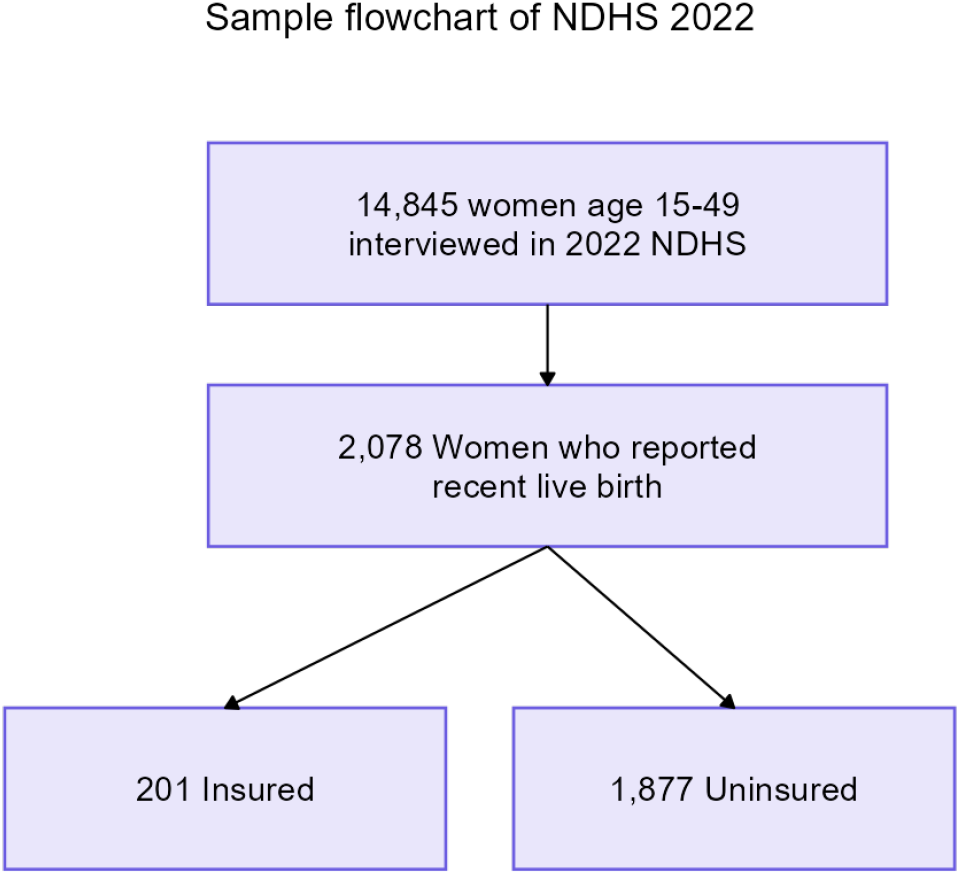
Flow chart of included study participants according to NDHS 2022 data.

#### Occupation

Occupation followed the DHS standard indicator, which records occupation only for women who were employed at some time in the 12 months preceding the survey (Ministry of Health and Population (Nepal) et al., 2023). White-collar comprises professional, technical and managerial, clerical, and sales and services occupations; blue-collar comprises skilled manual, unskilled manual, and paid domestic service work; agriculture comprises agricultural work. Homemakers and other women who were not employed in the previous 12 months are not assigned an occupation by the DHS, and are shown as “Not employed” in Table 1 rather than as a blue-collar or other occupational group.

**Table 1:** General characteristics of the study population, NDHS 2022.

| Respondent characteristics | Health insurance status |  |  | *p*-value<sup>2</sup> |
| --- | --- | --- | --- | --- |
| | Overall $N_w=1,998^1$ | No $N_w=1,778^1$ | Yes $N_w=220^1$ | |
| *Maternal age at delivery (years)* |  |  |  | <0.001*** |
| 15–24 | 1,011 (51%) | 940 (53%) | 71 (32%) |  |
| 25–34 | 876 (44%) | 742 (42%) | 135 (61%) |  |
| 35–49 | 111 (5.6%) | 96 (5.4%) | 14 (6.6%) |  |
| *Birth order* |  |  |  | <0.001*** |
| 1 | 811 (41%) | 686 (39%) | 125 (57%) |  |
| 2 | 703 (35%) | 631 (35%) | 72 (33%) |  |
| 3 | 291 (15%) | 276 (16%) | 15 (6.8%) |  |
| 4 or more | 193 (9.7%) | 185 (10%) | 8 (3.5%) |  |
| *Highest educational level* |  |  |  | <0.001*** |
| No Education | 370 (18%) | 363 (20%) | 7 (3.0%) |  |
| Primary | 674 (34%) | 638 (36%) | 37 (17%) |  |
| Secondary or Higher | 955 (48%) | 778 (44%) | 177 (80%) |  |
| *Wealth index* |  |  |  | <0.001*** |
| Poorest | 449 (22%) | 428 (24%) | 21 (9.7%) |  |
| Poorer | 439 (22%) | 413 (23%) | 26 (12%) |  |
| Middle | 397 (20%) | 363 (20%) | 34 (15%) |  |
| Richer | 400 (20%) | 349 (20%) | 51 (23%) |  |
| Richest | 314 (16%) | 225 (13%) | 88 (40%) |  |
| *Type of residence* |  |  |  | 0.009** |
| Urban | 1,307 (65%) | 1,141 (64%) | 166 (75%) |  |
| Rural | 692 (35%) | 638 (36%) | 54 (25%) |  |
| *Occupation* |  |  |  | <0.001*** |
| Agriculture | 902 (76%) | 815 (79%) | 87 (55%) |  |
| Blue-collar | 104 (8.8%) | 92 (8.9%) | 12 (7.8%) |  |
| White-collar | 177 (15%) | 119 (12%) | 58 (37%) |  |
| Not employed<sup>3</sup> | 816 | 752 | 64 |  |
| *Marital status* |  |  |  | >0.9 |
| Married or Living Together | 1,983 (99%) | 1,765 (99%) | 218 (99%) |  |
| Separated or Widowed | 15 (0.8%) | 14 (0.8%) | 2 (0.8%) |  |
| *Decision-making autonomy* |  |  |  | 0.014* |
| No | 1,332 (67%) | 1,206 (68%) | 126 (58%) |  |
| Yes | 651 (33%) | 559 (32%) | 93 (42%) |  |
| Unknown | 15 | 14 | 2 |  |
| *Problem accessing healthcare (distance)* |  |  |  | <0.001*** |
| No | 1,204 (60%) | 1,037 (58%) | 167 (76%) |  |
| Yes | 795 (40%) | 741 (42%) | 53 (24%) |  |

<sup>1</sup>The table includes all women who had given birth to a child within the two years preceding the survey. $N_w$ denotes a survey-weighted count; percentages are column percentages within each insurance status group. All counts and percentages are weighted by the NDHS sampling weights, so the column totals are 1,998 (1,778 uninsured, 220 insured) rather than the unweighted 2,078 (1,877 uninsured, 201 insured). No women were excluded. Because each cell is rounded separately, category counts may not sum exactly to the column totals.
<sup>2</sup>\* $p < 0.05$ ; \*\* $p < 0.01$ ; \*\*\* $p < 0.001$ ; chi-square tests with Rao–Scott adjustment for the survey design.
<sup>3</sup>Occupation percentages are calculated among women with an occupation recorded ( $N_w = 1,183$ weighted; 1,357 unweighted), that is, those employed in the previous 12 months; the 721 women (unweighted; 816 weighted) not employed in that period have no occupation recorded by the DHS. Decision-making autonomy percentages exclude the 12 women (unweighted; 15 weighted) with missing values, shown as “Unknown”; these were assigned the modal category for the propensity score model.

**Table 2:** Association between health insurance coverage and maternal health care utilisation.

| Characteristic | Health insurance status |  | AOR (95% CI)<sup>1</sup> |
| --- | --- | --- | --- |
|  | No (*N<sub>w</sub>*=1,778) | Yes (*N<sub>w</sub>*=220) |  |
| *Primary outcomes* |  |  |  |
| *ANC 4+ visits* |  |  |  |
| No | 364 (95%) | 17 (4.5%) | Ref |
| Yes | 1,370 (87%) | 201 (13%) | 2.14 (1.15 to 3.98) |
| *PNC check within two days* |  |  |  |
| No | 577 (96%) | 26 (4.4%) | Ref |
| Yes | 1,141 (86%) | 189 (14%) | 2.05 (1.28 to 3.27) |
| *PNC by skilled provider* |  |  |  |
| No | 590 (95%) | 28 (4.5%) | Ref |
| Yes | 1,127 (86%) | 187 (14%) | 1.94 (1.23 to 3.06) |
| *Secondary outcomes* |  |  |  |
| *Institutional delivery* |  |  |  |
| Non-institutional | 405 (96%) | 18 (4.3%) | Ref |
| Institutional | 1,373 (87%) | 202 (13%) | 1.55 (0.91 to 2.62) |
| *Skilled attendance* |  |  |  |
| Non-skilled | 381 (96%) | 17 (4.4%) | Ref |
| Skilled | 1,398 (87%) | 203 (13%) | 1.57 (0.90 to 2.72) |

<sup>1</sup>Cell entries are survey-weighted counts ( $N_w$ ) and row percentages; the unweighted analytic sample is 2,078 women (1,877 uninsured, 201 insured). Women with a missing outcome are excluded from that outcome's rows and model: 53 women (unweighted) for ANC4+ and 71 for each postnatal care outcome, so those rows sum to less than the column $N_w$ . The delivery outcomes are complete. AORs are from inverse probability of treatment weighted marginal structural models using the final analytical weight (survey weight × stabilised, truncated IPTW weight), with 95% confidence intervals from HC0 robust standard errors. Bootstrap intervals are given in Supplementary Table S5 and E-values in Supplementary Table S4.

Eight of these candidate confounders were included in the propensity score model: maternal age, birth order, educational attainment, household wealth status, marital status, place of residence, decision-making autonomy, and perceived difficulty in accessing healthcare due to distance. Three were identified in the DAG but excluded.

First, occupation is undefined for 34.7% of the sample because the DHS indicator is recorded only for women employed in the previous 12 months, so fitting it would have restricted the analysis to 1,357 women and conditioned the analysis on employment status. Second, province of residence is unevenly distributed with respect to the exposure: of the 422 women in Madhesh only 7 were insured, and 4 of the 7 provinces contributed fewer than 30 insured women. Its inclusion worsened overall covariate balance (maximum SMD 0.48 versus 0.16), reflecting limited overlap in insurance status across provinces. Third, recent internet use changed the estimated odds ratio by no more than 0.07. Supplementary Table S3 reports estimates with each variable restored.

This means that the propensity score model adjusts for socioeconomic position through education, wealth, and residence rather than through occupation and province. Because the excluded variables are strongly correlated with those retained, the practical consequence is limited; the sensitivity analyses in Supplementary Table S3 quantify it directly. The fitted model conditions on eight of the eleven confounders represented in the DAG. Therefore the model does not correspond to the minimal sufficient adjustment set implied by the graph. The reasons for each omission are given above (Tennant et al., 2021).

### 2.5 Statistical analysis

Descriptive statistics were used to summarise participant characteristics by insurance status. Categorical variables were presented as frequencies and percentages, and differences between insured and uninsured women were assessed using chi-square tests. Because health insurance enrolment is not randomly assigned, inverse probability of treatment weighting (IPTW) was used to reduce observed confounding and estimate the association between health insurance coverage and maternal healthcare utilisation. IPTW was selected instead of propensity score matching because it retains all observations while improving covariate balance between comparison groups and generating estimates that are more representative of the target population (Lunceford and Davidian, 2004; Austin and Stuart, 2015; King and Nielsen, 2019). A similar approach has been applied to Demographic and Health Survey data in comparable LMIC settings (Shifti et al., 2021).

Propensity scores were estimated using logistic regression, modelling the probability of health insurance enrolment as a function of the selected covariates. The propensity score model was specified as:

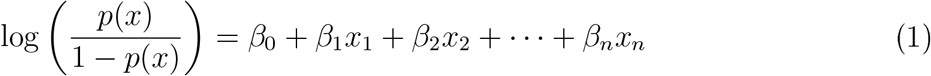

where *p*(*x*) represents the probability of health insurance enrolment conditional on the observed covariates and *β* represents the estimated regression coefficients (James et al., 2013).

Covariate balance before and after weighting was evaluated using the Standardised Mean Difference (SMD), which is less sensitive to sample size than significance testing (Austin and Stuart, 2015). The SMD for continuous variables was calculated as:

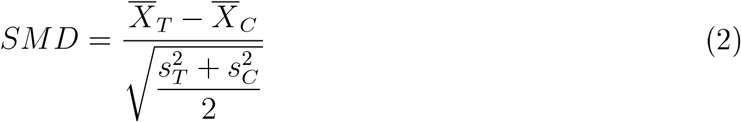

and for binary variables as:

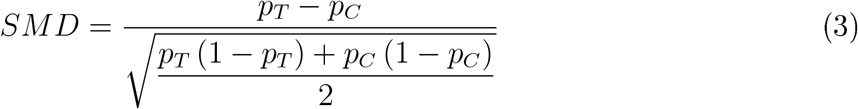

An SMD value below 0.10 was considered indicative of adequate balance between insured and uninsured groups (Cohen, 2013). Stabilised IPTW weights were calculated as follows (Cole and Hernán, 2008):

For insured participants:

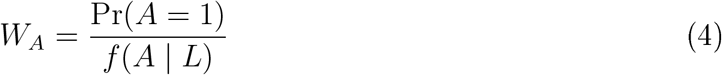

For uninsured participants:

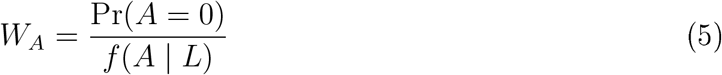

Weights were truncated at the 1st and 99th percentiles to minimise the influence of extreme values and improve model stability (Lee et al., 2011).

Because the NDHS employs a two-stage stratified cluster sampling design, survey weights were incorporated to obtain nationally representative estimates (Ministry of Health and Population (Nepal) et al., 2023). Final analytical weights were calculated by multiplying survey weights by stabilised IPTW weights, creating a weighted pseudo-population in which insurance status was independent of measured baseline covariates (DuGoff et al., 2014).

Three different denominators therefore appear in this paper and are labelled explicitly. The unweighted analytic sample is 2,078 women (201 insured, 1,877 uninsured); this is the figure reported in the participant flow diagram, and the number contributing to the propensity score model and to both delivery outcomes; the antenatal and postnatal models use fewer women because of missing outcome data (see “Missing data”). Tables 1 and 2 report survey-weighted counts and percentages, which sum to 1,998 (220 insured, 1,778 uninsured), because survey weights rescale the sample to the national population and insured women carry slightly larger weights on average. Supplementary Table S1 reports counts weighted by the stabilised IPTW weights alone, which sum to 2,063, since covariate balance is a property of the IPTW pseudo-population rather than of the survey-weighted population. No women were excluded between these three presentations.

Associations between health insurance coverage and maternal healthcare outcomes were estimated using marginal structural models (MSMs) of the form:

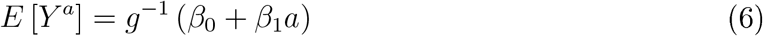

where *a* denotes health insurance exposure, *Y* represents the outcome of interest, and *g* is the logit link function (Robins et al., 2000). Results were reported as adjusted odds ratios (AORs) with 95% confidence intervals. Confidence intervals were estimated from heteroskedasticity-consistent (HC0) robust standard errors, which account for the weighting used in the marginal structural models. As a check on this choice, the models were additionally refitted using non-parametric bootstrap resampling with 1,000 iterations; the two approaches produced very similar intervals and identical conclusions for every outcome (Supplementary Table S5).

#### Missing data

Health insurance status and the two delivery outcomes (institutional delivery and skilled birth attendance) were complete for all 2,078 women. The DHS ANC4+ indicator was missing for 53 women (2.6%; 1 insured, 52 uninsured), and both postnatal care indicators were missing for 71 women (3.4%; 4 insured, 67 uninsured); 117 women (5.6%) lacked at least one primary outcome. Outcomes were not imputed: each marginal structural model was fitted to the women with that outcome observed (ANC4+, *n* = 2,025; each PNC outcome, *n* = 2,007; each delivery outcome, *n* = 2,078). Among the covariates entering the propensity score model, only decision-making autonomy contained missing values (12 women, 0.6%), which were assigned the modal category, so propensity scores and weights were estimated for all 2,078 women. Occupation was undefined for 721 women (34.7%) because they were not employed in the previous 12 months, as described above; it was not included in the propensity score model and was not imputed.

#### Sensitivity analyses

Three sets of sensitivity analyses were conducted. First, the propensity score model was refitted after adding each of the DAG covariates that had been left out of the primary specification, singly and jointly (Supplementary Table S3). Second, E-values were calculated to quantify how strong an unmeasured confounder would need to be, on the risk-ratio scale and conditional on the measured covariates, to explain away the observed associations (VanderWeele and Ding, 2017). Because the outcomes are common, odds ratios were converted to approximate risk ratios as 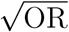 before computing E-values (Supplementary Table S4). Third, the bootstrap comparison described above was performed (Supplementary Table S5).

All analyses were conducted using R, including the survey package (Lumley, 2024), and tables were generated with the gtsummary package (Sjoberg et al., 2021).

## 3 Results

### 3.1 Participant characteristics

The 2022 NDHS interviewed 14,845 women aged 15–49 years. For this analysis, women who reported a live birth within the 24 months preceding the survey were included, resulting in a final analytical sample of 2,078 women. Of these, 201 (9.7%) reported health insurance coverage, while 1,877 (90.3%) reported no insurance coverage, as shown in Figure 2. Decision-making autonomy was missing for 12 women (0.6%) and was assigned the modal category. ANC4+ was missing for 53 women and the two postnatal care outcomes for 71 women, who were excluded from the models for those outcomes only (see Methods, “Missing data”). Counts in Tables 1 and 2 are survey-weighted and sum to 1,998, as set out in the Methods.

Table 1 presents the sociodemographic characteristics of study participants according to health insurance status. Insurance coverage varied significantly across several demographic and socioeconomic characteristics. Women aged 25–34 years were more likely to be insured than younger women aged 15–24 years (*p <* 0.001). Insurance coverage was also higher among women with their first birth, those who had completed secondary or higher education, women in the richest wealth quintile, white-collar workers, and urban residents (all *p <* 0.05). Recent internet use, which is not tabulated in Table 1, showed the same pattern: 83.0% of insured women had used the internet in the previous 12 months compared with 63.7% of uninsured women (*p <* 0.001). Conversely, insurance enrolment was substantially lower among women with no formal education, those in poorer wealth quintiles, women with higher birth orders, agricultural workers, and rural residents.

Women in the richest wealth quintile were approximately six times more likely to be insured than women in the poorest quintile (28% vs 4.7%), and women with secondary or higher education accounted for 80% of insured women. These findings suggest that women who are already socioeconomically advantaged are more likely to benefit from financial protection mechanisms during pregnancy and childbirth.

### 3.2 Association between health insurance and maternal healthcare utilisation

After adjustment for observed confounding using IPTW, health insurance coverage was positively associated with utilisation of antenatal and postnatal healthcare services (Table 2). Women with health insurance had more than twice the odds of attending at least four ANC visits compared with uninsured women (AOR 2.14; 95% CI 1.15–3.98). Attainment of the WHO-recommended eight antenatal contacts was rare. Among women with a recorded number of ANC visits, 80.5% attended at least four but only 6.0% attended eight or more, and 42.5% reported exactly four, consistent with care being anchored to the nationally incentivised four-visit target. All three figures are survey-weighted. Similarly, insured women had approximately twice the odds of receiving a postnatal check-up within two days of delivery (AOR 2.05; 95% CI 1.28–3.27). Insurance coverage was also associated with a significantly higher likelihood of receiving PNC from a skilled provider (AOR 1.94; 95% CI 1.23–3.06). For delivery-related outcomes, the direction of association remained positive but statistical evidence was weaker. Insured women had higher odds of delivering in a health facility (AOR 1.55; 95% CI 0.91–2.62) and receiving skilled birth attendance (AOR 1.57; 95% CI 0.90–2.72); however, neither association reached statistical significance. Figure 3 summarises these estimates.

**Figure 3:**
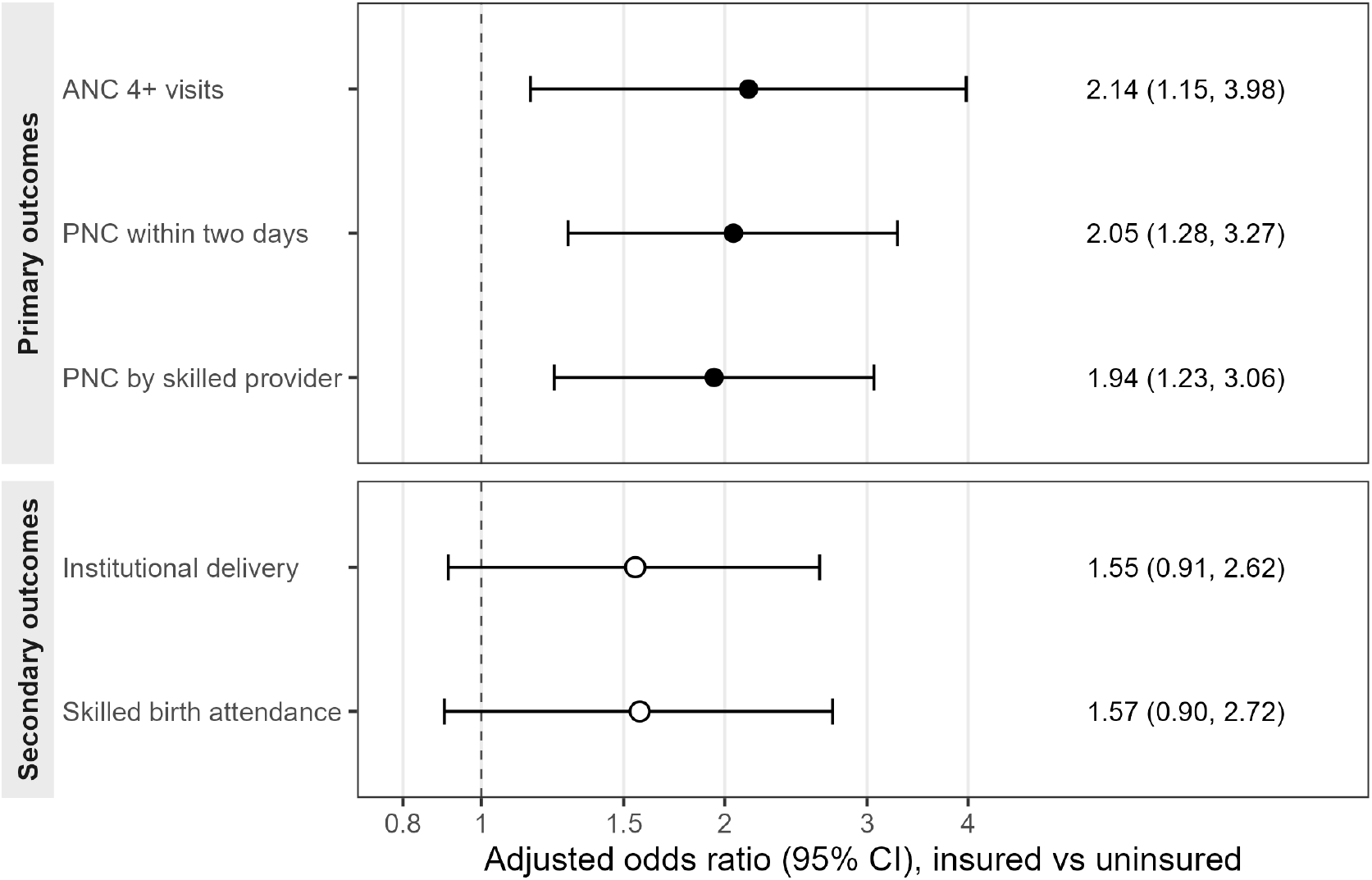
Adjusted odds ratios for maternal healthcare utilisation by health insurance status. Estimates are those reported in Table 2.

The IPTW model demonstrated satisfactory performance. Most post-weighting SMDs were below 0.10, indicating adequate covariate balance between insured and uninsured groups. Small residual imbalances remained for age and educational attainment, but these were substantially reduced compared with the unweighted data and are within acceptable limits for observational analyses. In addition, examination of stabilised weights suggested no major violations of the positivity assumption. Detailed covariate balance and positivity assessment results are presented in Supplementary Tables S1–S2 and Supplementary Figure S1.

### 3.3 Sensitivity analyses

The associations for the postnatal outcomes were stable across alternative propensity score specifications (Supplementary Table S3). Adding province of residence left the postnatal and delivery estimates close to the primary specification and did not alter any conclusion, although the ANC4+ estimate rose somewhat (AOR 2.46, 95% CI 1.28–28.73; PNC within two days AOR 1.98, 95% CI 1.22–22.23) and overall covariate balance deteriorated markedly (maximum SMD 0.476 versus 0.156). Adding recent internet use changed no estimate materially. Restricting the model to the 1,357 women for whom occupation is defined attenuated the estimates and widened the intervals: the ANC4+ association was no longer statistically significant (AOR 1.83; 95% CI 0.86–86.89) and the secondary delivery outcomes moved towards the null (institutional delivery AOR 1.02; 95% CI 0.60–60.75). The two PNC associations remained positive and, for PNC within two days, statistically significant (AOR 1.75; 95% CI 1.04–04.95). Because this restriction removes all women not employed in the previous 12 months rather than a random subset, the attenuation reflects both the loss of a third of the sample and a change in the population being described, and it should not be read as evidence that the primary estimates are confounded by occupation.

E-values indicated moderate sensitivity to unmeasured confounding (Supplementary Table S4). For the ANC4+ association, an unmeasured confounder would need to be associated with both insurance enrolment and ANC4+ attendance by a risk ratio of at least 2.29, above and beyond the measured covariates, to explain away the point estimate, and at least 1.35 to shift the confidence interval to include the null. Corresponding E-values were 2.22 and 1.52 for PNC within two days, and 2.13 and 1.46 for PNC from a skilled provider. Bootstrap confidence intervals based on 1,000 resamples were very close to the reported robust intervals and did not alter any conclusion (Supplementary Table S5).

## 4 Discussion

This study examined the association between health insurance coverage and maternal healthcare utilisation among women with a recent live birth in Nepal using nationally representative NDHS 2022 data and IPTW. While insurance coverage was generally low, the findings indicate that health insurance coverage was associated with substantially greater utilisation of antenatal and postnatal healthcare services. Insured women had more than twice the odds of attending at least four ANC visits and receiving a PNC check-up within two days of delivery, while also being significantly more likely to receive PNC from a skilled provider. Although insured women had higher odds of institutional delivery and skilled birth attendance, these associations did not reach statistical significance. These findings suggest that health insurance may play an important role in facilitating maternal healthcare utilisation, particularly for preventive and follow-up services, while financial protection alone may be insufficient to overcome barriers influencing delivery care utilisation. Because the NDHS is cross-sectional and enrolment is voluntary, these estimates describe differences between insured and uninsured women after balancing measured covariates, not the effect of enrolling a woman in the insurance scheme.

The observed positive association between health insurance coverage and ANC utilisation is consistent with evidence from Nepal and other LMICs. Analyses of successive Nepal Demographic and Health Surveys have documented persistent socioeconomic gradients in ANC utilisation and incomplete coverage across the maternal continuum of care (Khatri et al., 2025; Pandey et al., 2025). Previous analyses from Ghana, Indonesia, Rwanda, and multi-country Demographic and Health Survey datasets have shown that insured women are more likely to initiate ANC and complete recommended visits than uninsured women (Browne et al., 2016; Wang et al., 2017). Health insurance reduces direct out-of-pocket expenditures and the uncertainty associated with healthcare costs, which may encourage women to seek preventive care earlier and more regularly throughout pregnancy. In Nepal, despite considerable progress in maternal health service coverage, financial barriers remain an important determinant of healthcare utilisation, particularly among poorer households and those residing in rural areas. The findings therefore support ongoing efforts to strengthen financial protection mechanisms as part of broader strategies to improve maternal health outcomes and progress towards universal health coverage.

The gap between the two ANC thresholds is striking: 80.5% of women attended at least four visits, but only 6.0% attended eight or more, and 42.5% reported exactly four. This pattern is consistent with national policy shaping care-seeking. Four visits is the standard the Aama Programme incentivises and against which performance is monitored, whereas the eight-contact model has been endorsed but not rolled out (Ministry of Health and Population (Nepal), 2019). Coverage appears to track the incentivised target rather than the aspirational one, which suggests that meeting the Road Map’s commitment to eight contacts will require more than financial protection alone.

The strong association between insurance coverage and timely PNC utilisation is particularly noteworthy. Although substantial improvements have been made in maternal health service coverage in Nepal, PNC remains one of the weakest links within the maternal and newborn continuum of care. Previous studies have consistently reported lower coverage of timely PNC compared with ANC and facility delivery services (Pandey et al., 2025; Thapa et al., 2024). The present study suggests that financial protection provided through health insurance may facilitate continued engagement with healthcare services beyond childbirth, reducing barriers to postnatal follow-up and increasing access to skilled providers.

This is notable because the existing evidence base for an effect of insurance on PNC is limited. A systematic review of low- and lower-middle-income countries found positive effects on ANC and delivery care but no evidence of an effect on PNC utilisation (Kazibwe et al., 2024). The present finding therefore adds to a relatively sparse literature rather than confirming an established pattern. Furthermore, by using nationally representative NDHS 2022 data and propensity score weighting to balance measured confounders, this study provides updated evidence from Nepal that complements and extends findings from earlier studies conducted using older datasets.

In contrast, health insurance coverage was not significantly associated with institutional delivery or skilled birth attendance after adjustment for confounding factors. This differs from pooled evidence across low- and lower-middle-income countries, where insurance has generally shown positive effects on facility delivery and skilled attendance (Kazibwe et al., 2024).

A likely explanation lies in how delivery care is financed in Nepal. Since 2009, the Aama Programme has removed user fees for all types of institutional delivery and provides a cash transport incentive paid immediately after facility-based birth (Ministry of Health and Population (Nepal), 2019). Delivery care is therefore already free at the point of use for every woman, insured or not, whereas ANC and PNC are not subsidised in the same way. The marginal financial benefit that insurance confers is consequently smallest precisely for the delivery-related outcomes, which may explain why the associations observed here are strongest for antenatal and postnatal services and weakest for institutional delivery and skilled birth attendance. Whether delivery care is free in practice is a separate question: informal payments, medicines, and ancillary costs may persist despite the formal fee exemption, and the NDHS does not capture out-of-pocket expenditure.

A further explanation is that decisions regarding place of delivery are influenced by a wider range of factors beyond affordability. Geographic accessibility, transportation challenges, cultural preferences, family decision-making, perceptions of quality of care, and health system readiness all contribute to whether women deliver in health facilities (Thapa et al., 2023; Thasineku et al., 2025). In Nepal’s mountainous and rural settings, these structural and sociocultural barriers may continue to constrain facility delivery even when financial barriers are reduced through insurance coverage. Consequently, improving institutional delivery rates is likely to require coordinated interventions that combine financial protection with strengthened healthcare infrastructure, improved referral systems, transportation support, and community engagement. Most of these constraints apply across the continuum of care. Delivery differs in that labour is unpredictable and cannot be deferred: it may begin at night or during the monsoon, requires immediate transport, and depends on a facility being staffed at that moment. Insurance does not alter these features.

An important finding of this study relates to inequities in health insurance coverage. Insurance enrolment was substantially higher among women who were socioeconomically advantaged, including those with higher levels of education, greater household wealth, urban residence, white-collar occupations, and internet access. Women in the richest wealth quintile were considerably more likely to be insured than women in the poorest quintile, suggesting that financial protection mechanisms may not yet be reaching the populations most vulnerable to adverse maternal health outcomes. Similar inequities in insurance enrolment and maternal healthcare utilisation have been reported previously in Nepal and other LMICs (Anindya et al., 2020; Khatri et al., 2025). This pattern raises an important policy concern. If health insurance primarily benefits women who are already relatively advantaged, expansion of insurance coverage could inadvertently widen existing inequalities in maternal health service utilisation. Strengthening equity-oriented enrolment strategies, targeting vulnerable populations, and routinely monitoring coverage across socioeconomic groups should therefore be prioritised as Nepal expands its health insurance programme.

The findings should also be interpreted in the context of Nepal’s persistently low health insurance coverage. According to the 2022 NDHS, most women and men remain uninsured despite the expansion of the National Health Insurance Program. Previous research has identified multiple factors contributing to low enrolment and high dropout rates, including limited awareness of programme benefits, perceived deficiencies in service quality, administrative challenges, and dissatisfaction with benefit packages (Khanal et al., 2023). The positive associations observed in this study suggest that increasing enrolment and retention within the insurance system could contribute to improvements in maternal healthcare utilisation. However, expanding coverage alone is unlikely to be sufficient. Policymakers should also address concerns regarding quality of care, service availability, and user experience to ensure that insurance translates into meaningful health gains.

This study has several strengths. First, it utilised nationally representative data from the most recent NDHS, thereby providing findings that are broadly generalisable to women of reproductive age in Nepal. Second, the application of a DAG informed the selection of confounders, reducing the risk of inappropriate adjustment. Third, IPTW allowed the creation of a weighted pseudo-population in which measured baseline characteristics were balanced between insured and uninsured women, strengthening the validity of estimated associations. Assessment of SMDs and positivity assumptions indicated satisfactory covariate balance and support for the weighting approach.

Several limitations should also be considered. First, the cross-sectional nature of the NDHS precludes definitive causal inference, and the findings should therefore be interpreted as associations rather than causal effects. Second, although IPTW reduces bias arising from measured confounding, residual confounding due to unobserved factors cannot be excluded. Factors such as health-seeking behaviour, perceptions of healthcare quality, social support, and detailed household characteristics may influence both insurance enrolment and maternal healthcare utilisation, and enrolment in the NHIP is voluntary, so women who enrol may differ systematically from those who do not in ways the NDHS does not measure. The E-values bound this concern rather than resolve it: an unmeasured confounder would need a risk ratio of about 2.3 with both enrolment and ANC4+ attendance to explain away the strongest association, but only about 1.35 to render its confidence interval compatible with the null. Health-seeking tendency could plausibly reach that magnitude. These associations are consistent with, but do not demonstrate, a benefit of insurance.

Third, the propensity score model adjusted for eight of the eleven confounders in the DAG. Occupation was excluded because including it would restrict the analysis to employed women, and province could not be balanced because insured women were unevenly distributed across provinces. Therefore, socioeconomic position was captured through education, wealth, and residence. Supplementary Table S3 shows that adding province does not materially change the estimates, while restricting to employed women weakens the antenatal and delivery associations through a change in study population rather than through confounding by occupation. An exploratory analysis using an eight-visit threshold gave an adjusted odds ratio of 1.89 (95% CI 1.00–00.57), directionally consistent with the ANC4+ association but based on only 21 insured women achieving eight visits, and unstable across alternative propensity score specifications. This threshold was therefore not reported as an outcome. The concentration of responses at exactly four visits also limits the interpretability of higher thresholds, and the DHS records visits rather than the contacts specified in the WHO model. Fourth, health insurance was measured as a binary exposure because it is overwhelmingly a government-scheme measure: 91.2% of insured women were enrolled in government, social security or employer schemes, 7.3% held private commercial cover only, and 1.4% held both. Differences in utilisation between specific schemes could therefore not be examined. Fifth, insurance status was recorded at the survey, not during pregnancy. Thus, for some women, enrolment may have occurred after the care was received, and the direction of this bias is unknown. Finally, maternal healthcare utilisation was self-reported and may therefore be subject to recall bias, although the analysis was restricted to births occurring within the previous 24 months to minimise this concern.

## 5 Conclusion

Health insurance coverage was positively associated with utilisation of antenatal and post-natal healthcare services among women in Nepal. Insured women were significantly more likely to complete recommended ANC visits and receive timely and skilled PNC. However, no significant association was observed for institutional delivery or skilled birth attendance, suggesting that financial protection alone may not overcome the full range of barriers affecting maternal healthcare utilisation. The study also identified substantial inequities in insurance enrolment, with coverage concentrated among socioeconomically advantaged women. Efforts to expand health insurance coverage should therefore be accompanied by targeted strategies to improve enrolment among vulnerable populations and by broader investments in health system strengthening. Together, these measures may contribute to improving maternal health outcomes and supporting Nepal’s progress towards universal health coverage and the SDGs.

## Data Availability

The data analysed in this study are publicly available from The DHS Program. The 2022 Nepal Demographic and Health Survey datasets can be obtained free of charge at https://www.dhsprogram.com following registration and approval of a data request. The authors had no special access privileges. Analysis code is available from the corresponding author on request.

https://dhsprogram.com/data/dataset/Nepal_Standard-DHS_2022.cfm

## Author Contributions

**Yash Raj Lamsal:** Conceptualisation, Methodology, Software, Formal analysis, Data curation, Visualisation, Writing - original draft, Writing - review & editing, Project administration. **Abdulaziz Mohammed Hussen:** Methodology, Validation, Writing - review & editing. **Joyce L. Browne:** Supervision, Writing - review & editing. **Reshu Agrawal Sagtani:** Writing - original draft, Validation, Supervision, Writing - review & editing. All authors read and approved the final manuscript.

## Data Availability

NDHS 2022 data are publicly available from the DHS Program at https://www.dhsprogram.comdhsprogran con upon registration.

## Acknowledgements

The authors acknowledge the DHS Program and the Ministry of Health and Population of Nepal for providing access to the NDHS 2022 data. The authors thank the survey respondents and field teams of the NDHS 2022.

## Ethics Approval

The survey protocol for the NDHS 2022 was reviewed by the Nepal Health Research Council and the ICF Institutional Review Board, and written informed consent was obtained from all respondents. The authors obtained permission to use the data from the DHS Program. The study used de-identified, publicly available data, and no additional ethical approval was required.

## Use of AI tools

Generative AI tools were used during code development and manuscript preparation. Claude (Anthropic) assisted with R code development, LaTeX formatting, and prose drafting and editing; Grammarly was used by individual authors for language refinement. AI tools were not used for data collection, statistical analysis, interpretation of results, or the generation of scientific conclusions, and were not used to generate or modify any primary data. All AI-assisted code was verified by re-running the analysis pipeline and confirming that every reported estimate reproduced exactly; all references were independently checked against the published sources. All hypotheses, interpretations, results, conclusions, limitations and implications reported here are the authors’ own. The authors take full responsibility for the content of the manuscript.

## Supplementary Information

### Covariate balance assessment

To assess the inverse probability of treatment weighting (IPTW) approach, covariate balance between insured and uninsured women was evaluated using standardised mean differences (SMDs) before and after weighting. As shown in Supplementary Table S1, weighting substantially improved the balance of characteristics between treated and untreated groups. For most covariates the SMD was below the recommended threshold of 0.10 following weighting, indicating adequate comparability between insured and uninsured women. Small residual imbalances remained for age and educational attainment, but these values were substantially reduced compared with the unweighted data and are within acceptable limits for observational analyses. Overall, the results suggest that the IPTW procedure effectively reduced observed confounding and improved exchangeability.

### Propensity score overlap

The distribution of propensity scores was examined to assess overlap between insured and uninsured women. In Supplementary Figure S1, there was substantial overlap in propensity score distributions between exposure groups after weighting. The weighted density plots demonstrate a better alignment between the two groups, indicating that women with similar observed characteristics had a non-zero probability of both receiving and not receiving health insurance coverage. These findings provide support for the positivity assumption required for valid IPTW estimation.

### Positivity assessment

The positivity assumption was further evaluated using the distribution of stabilised and unstabilised weights (Supplementary Table S2). The mean stabilised weight was 0.99, close to the expected value of 1.0, while the mean unstabilised weight was 2.10. Stabilised weights ranged from 0.22 to 2.52, with no extreme values. This suggests that weight stabilisation and truncation adequately limited the influence of extreme observations. Overall, there was no major violation of the positivity assumption, supporting the IPTW analysis.

### Alternative propensity score specifications

The primary propensity score model included eight of the eleven confounders in the DAG. Supplementary Table S3 shows the results from adding excluded covariates. Adding recent internet use has small effects on the estimates. Adding province of residence leaves the conclusions unchanged but raises the ANC4+ estimate from 2.14 to 2.46 and worsens overall covariate balance (maximum SMD 0.476 versus 0.156) because insured women were distributed unevenly across provinces; that deterioration in balance is why province was not carried into the primary specification. Adding occupation restricted the analysis to only 1,357 employed women because occupation was not available for the remaining women in the DHS. This restriction reduced the antenatal and delivery-related estimates. Thus, this comparison reflects a change in the study population rather than confounding by occupation.

**Table S1:** Standardised mean differences (SMDs) for baseline covariates after IPTW among women with a live birth in the preceding 24 months, NDHS 2022.

| **Covariate** | **Uninsured $n_w$ (%)** | **Insured $n_w$ (%)** | **SMD** |
| --- | --- | --- | --- |
| *Maternal age* |  |  |  |
| 15–24 years | 1001.7 (53.2) | 91.2 (50.2) | 0.156 |
| 25–34 years | 775.7 (41.2) | 73.1 (40.2) |  |
| 35–49 years | 103.8 (5.5) | 17.5 (9.6) |  |
| *Birth order* |  |  |  |
| 1 | 773.7 (41.1) | 78.2 (43.0) | 0.099 |
| 2 | 643.9 (34.2) | 55.9 (30.8) |  |
| 3 | 264.8 (14.1) | 30.3 (16.7) |  |
| 4 or more | 198.9 (10.6) | 17.4 (9.6) |  |
| *Education status* |  |  |  |
| No Education | 331.2 (17.6) | 22.6 (12.5) | 0.146 |
| Primary | 649.7 (34.5) | 65.2 (35.9) |  |
| Secondary or Higher | 900.3 (47.9) | 94.0 (51.7) |  |
| *Wealth index* |  |  |  |
| Poorest | 617.5 (32.8) | 61.8 (34.0) | 0.044 |
| Poorer | 407.3 (21.7) | 36.8 (20.2) |  |
| Middle | 331.5 (17.6) | 33.1 (18.2) |  |
| Richer | 315.5 (16.8) | 31.0 (17.1) |  |
| Richest | 209.4 (11.1) | 19.1 (10.5) |  |
| Marital status (separated/widowed) | 11.2 (0.6) | 1.6 (0.9) | 0.031 |
| Rural residence | 917.1 (48.8) | 84.0 (46.2) | 0.051 |
| Decision-making autonomy | 623.6 (33.1) | 60.7 (33.4) | 0.006 |
| Distance to health facility | 829.7 (44.1) | 85.7 (47.1) | 0.061 |

**Table S2:**
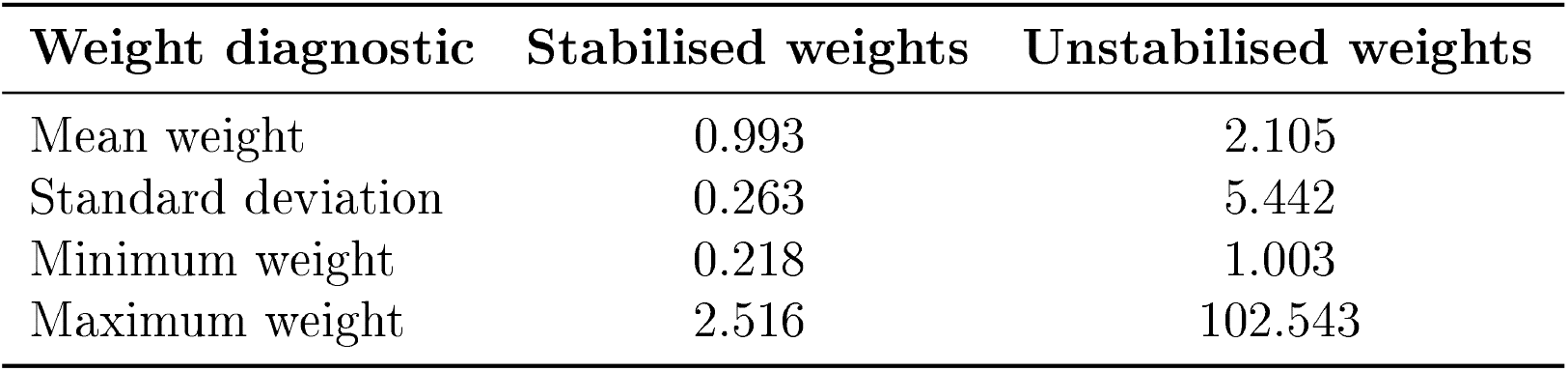
Positivity diagnostics and weight distribution used in the analysis, NDHS 2022.

**Figure S1:**
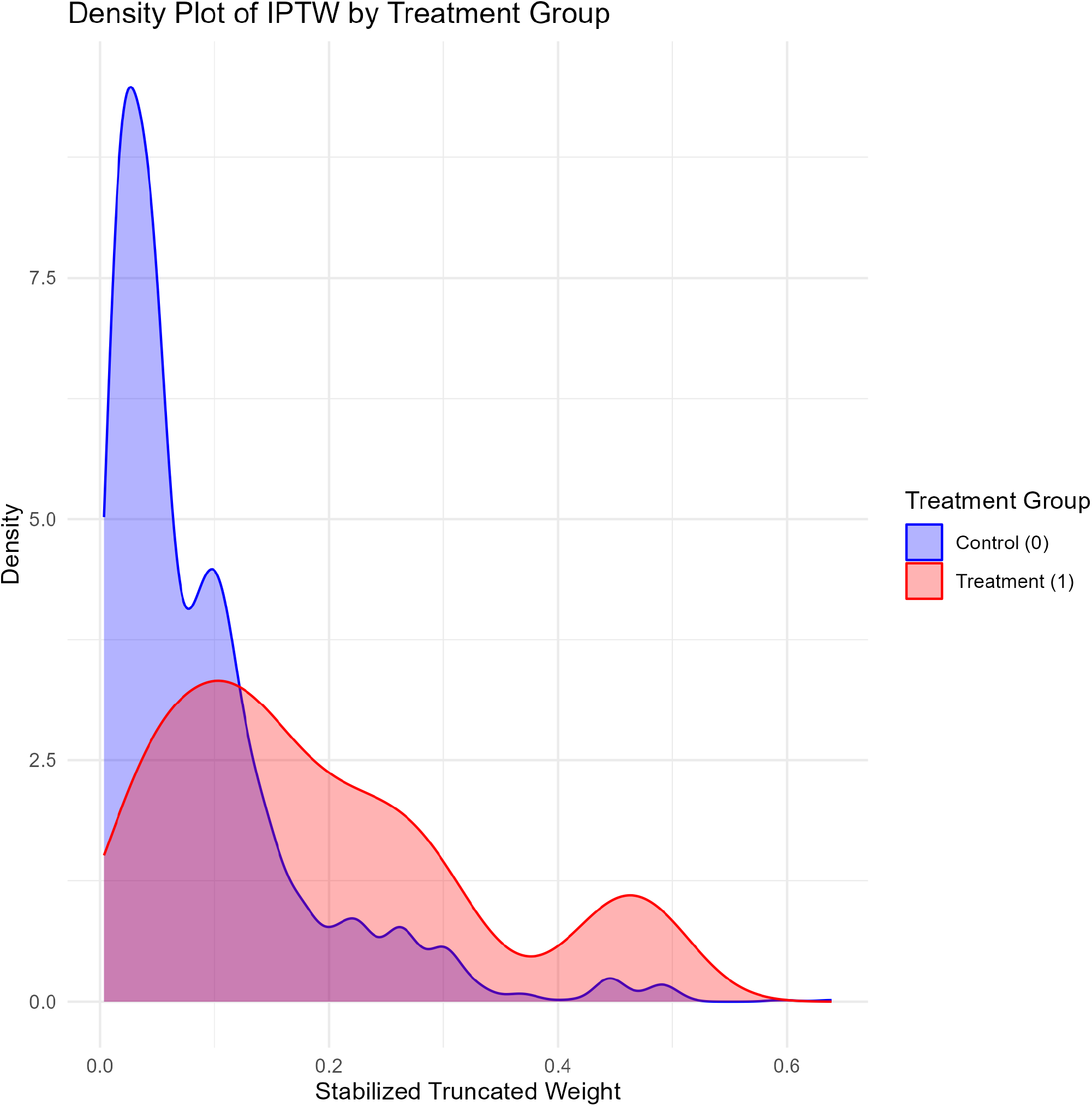
Propensity score distribution and weighted density plots.

**Table S3:** Adjusted odds ratios under alternative propensity score specifications, NDHS 2022.

| Propensity score model | *n* | ANC 4+ | PNC $\leq 2$ d | PNC skilled | Inst. deliv. | SBA | Max SMD |
| --- | --- | --- | --- | --- | --- | --- | --- |
| Primary analysis (Table 2) | 2,078 | 2.14<br>(1.15–3.98) | 2.05<br>(1.28–3.27) | 1.94<br>(1.23–3.06) | 1.55<br>(0.91–2.62) | 1.57<br>(0.90–2.72) | 0.156 |
| + province of residence | 2,078 | 2.46<br>(1.28–4.73) | 1.98<br>(1.22–3.23) | 1.91<br>(1.19–3.07) | 1.59<br>(0.91–2.76) | 1.59<br>(0.89–2.83) | 0.476 |
| + recent internet use | 2,078 | 2.07<br>(1.11–3.85) | 2.03<br>(1.27–3.26) | 1.93<br>(1.22–3.05) | 1.52<br>(0.89–2.57) | 1.53<br>(0.88–2.66) | 0.160 |
| + occupation (employed women only) | 1,357 | 1.83<br>(0.86–3.89) | 1.75<br>(1.04–2.95) | 1.59<br>(0.96–2.63) | 1.02<br>(0.60–1.75) | 1.13<br>(0.64–1.99) | 0.195 |
| + province, occupation, internet use | 1,357 | 1.84<br>(0.83–4.08) | 1.87<br>(1.11–3.15) | 1.70<br>(1.02–2.82) | 1.07<br>(0.62–1.85) | 1.19<br>(0.67–2.11) | 0.340 |

Values are adjusted odds ratios with 95% confidence intervals from HC0 robust standard errors. Each row refits the entire weighting procedure — propensity score model, stabilised weights, truncation at the 1st and 99th percentiles, and multiplication by the survey weight — under the stated covariate set. “Max SMD” is the largest post-weighting standardised mean difference across the covariates in that model; values further from zero indicate poorer balance. *n* is the unweighted number of women contributing to the model.

### E-values for unmeasured confounding

Because health-seeking behaviour may affect both insurance enrolment and healthcare use, we calculated E-values to assess the potential impact of unmeasured confounding (VanderWeele and Ding, 2017). Odds ratios were converted to approximate risk ratios using 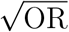. An E-value of 2.29 for the ANC4+ association means that an unmeasured confounder would need to be associated with both insurance enrolment and ANC4+ attendance by a risk ratio of at least 2.29 each, conditional on the measured covariates, to reduce the observed association to the null. Weaker confounding of at least 1.35 would suffice to move the confidence interval to include the null. These are moderate rather than large values, and they do not rule out confounding by an unmeasured health-seeking disposition.

**Table S4:** E-values for the reported associations, NDHS 2022.

| Outcome | AOR (95% CI) | E-value, estimate | E-value, CI |
| --- | --- | --- | --- |
| ANC 4+ visits | 2.14 (1.15–3.98) | 2.29 | 1.35 |
| PNC within 2 days | 2.05 (1.28–3.27) | 2.22 | 1.52 |
| PNC by skilled provider | 1.94 (1.23–3.06) | 2.13 | 1.46 |
| Institutional delivery | 1.55 (0.91–2.62) | 1.80 | 1.00 |
| Skilled birth attendance | 1.57 (0.90–2.72) | 1.82 | 1.00 |

An E-value of 1.00 for the confidence interval indicates that the interval already includes the null, so no unmeasured confounding is required to make the association compatible with no difference.

### Comparison of robust and bootstrap confidence intervals

Table S5 compares confidence intervals based on HC0 robust standard errors in Table 2 with percentile intervals from 1,000 bootstrap samples. The bootstrap means exceed the corresponding point estimates because the sampling distribution of an odds ratio is right-skewed, and the intervals are wider, most noticeably for ANC4+ (1.23–23.77 versus 1.15–15.98). The two approaches agree on statistical significance for every outcome.

**Table S5:** HC0 robust versus bootstrap confidence intervals, NDHS 2022.

| Outcome | Robust (HC0), as reported | Bootstrap, 1,000 resamples |
| --- | --- | --- |
| ANC 4+ visits | 2.14 (1.15–3.98) | 2.39 (1.23–4.77) |
| PNC within 2 days | 2.05 (1.28–3.27) | 2.14 (1.36–3.37) |
| PNC by skilled provider | 1.94 (1.23–3.06) | 2.02 (1.30–3.15) |
| Institutional delivery | 1.55 (0.91–2.62) | 1.65 (0.97–2.80) |
| Skilled birth attendance | 1.57 (0.90–2.72) | 1.69 (0.96–2.96) |

Bootstrap entries are the mean of the resampled adjusted odds ratios with 2.5th and 97.5th percentile limits. Resampling was performed on the weighted analytic dataset, holding the estimated weights fixed.

### STROBE checklist

**Table S6:**
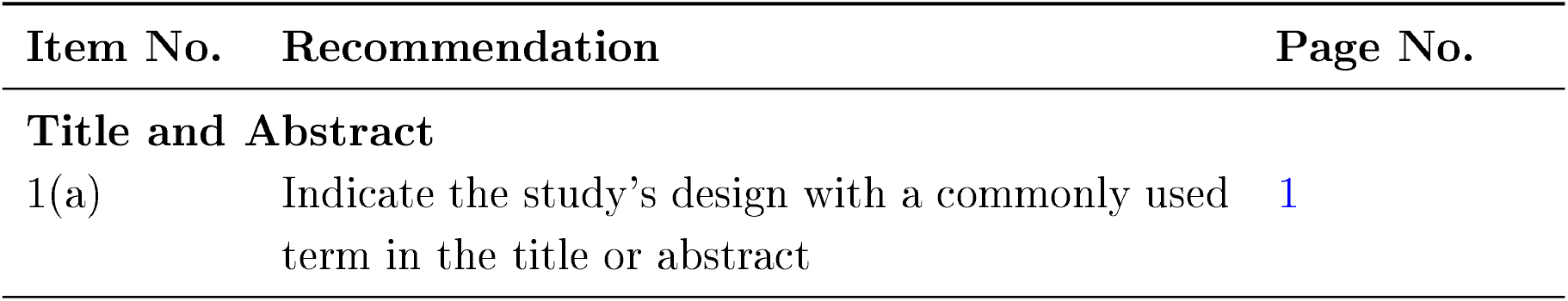
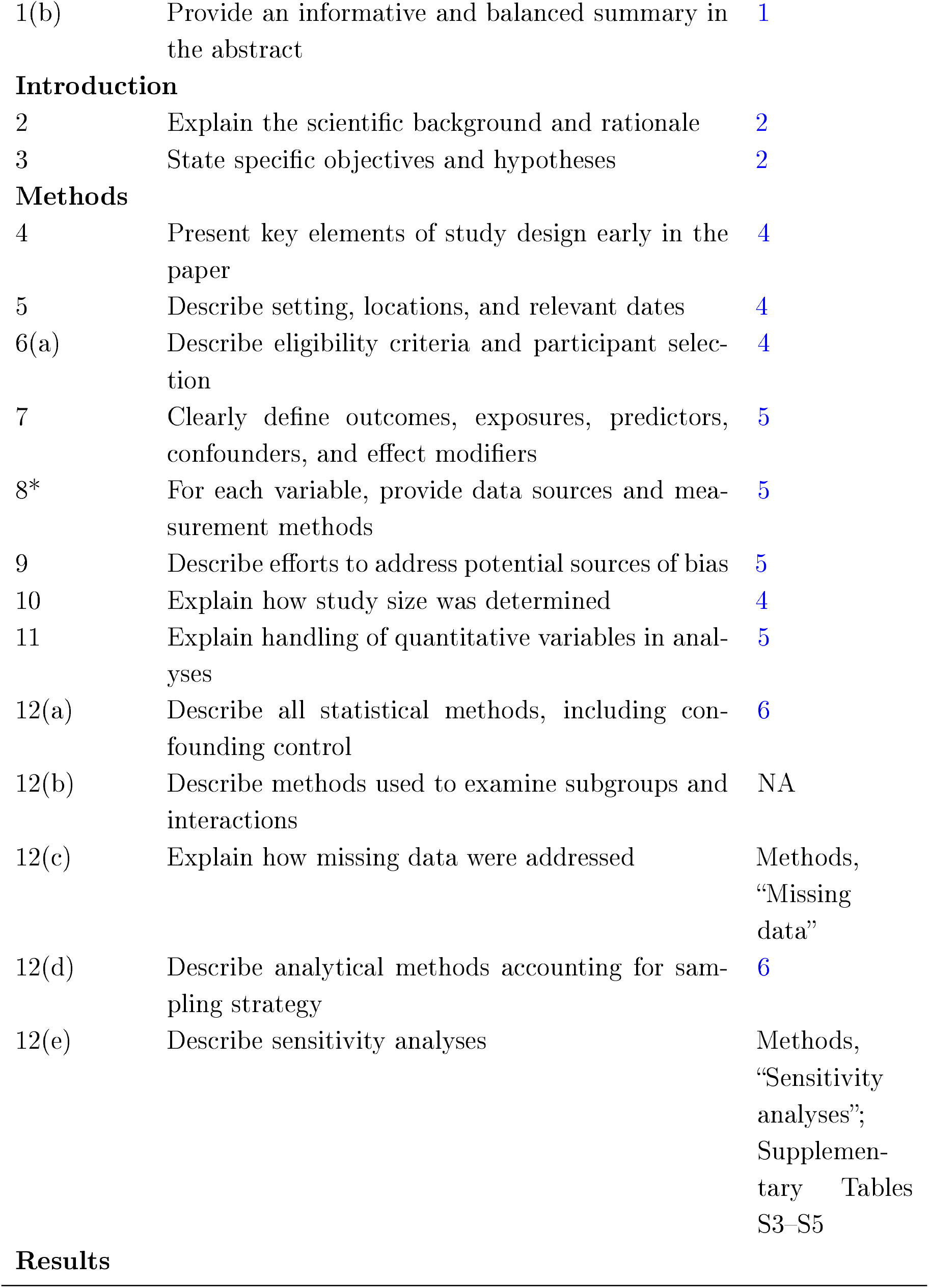
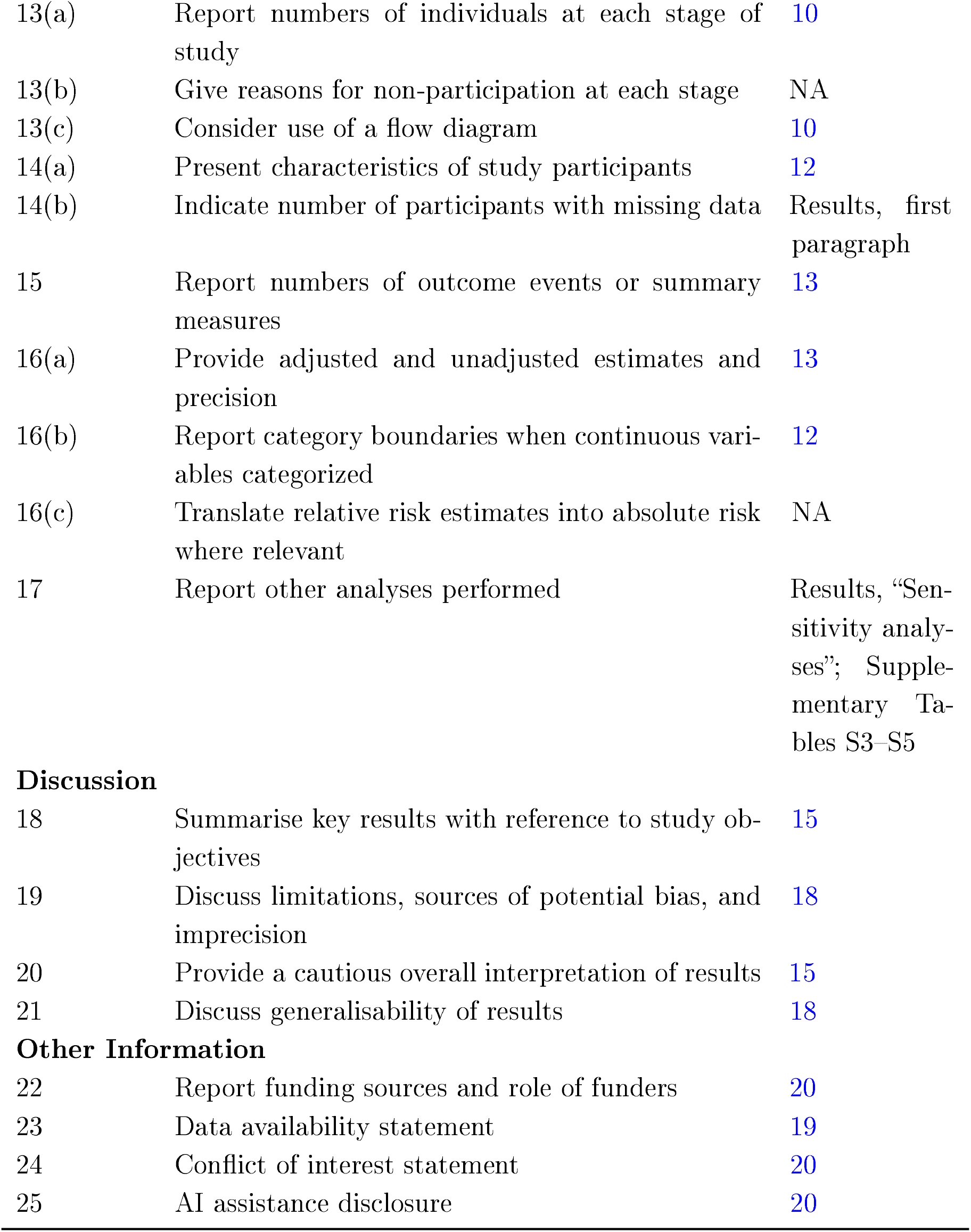
STROBE checklist for cross-sectional studies.

| Item No. | Recommendation | Page No. |
| --- | --- | --- |
| **Title and Abstract** |  |  |
| 1(a) | Indicate the study’s design with a commonly used term in the title or abstract | 1 |
| 1(b) | Provide an informative and balanced summary in the abstract | 1 |
| **Introduction** |  |  |
| 2 | Explain the scientific background and rationale | 2 |
| 3 | State specific objectives and hypotheses | 2 |
| **Methods** |  |  |
| 4 | Present key elements of study design early in the paper | 4 |
| 5 | Describe setting, locations, and relevant dates | 4 |
| 6(a) | Describe eligibility criteria and participant selection | 4 |
| 7 | Clearly define outcomes, exposures, predictors, confounders, and effect modifiers | 5 |
| 8* | For each variable, provide data sources and measurement methods | 5 |
| 9 | Describe efforts to address potential sources of bias | 5 |
| 10 | Explain how study size was determined | 4 |
| 11 | Explain handling of quantitative variables in analyses | 5 |
| 12(a) | Describe all statistical methods, including confounding control | 6 |
| 12(b) | Describe methods used to examine subgroups and interactions | NA |
| 12(c) | Explain how missing data were addressed | Methods, “Missing data” |
| 12(d) | Describe analytical methods accounting for sampling strategy | 6 |
| 12(e) | Describe sensitivity analyses | Methods, “Sensitivity analyses”; Supplementary Tables S3–S5 |
| **Results** |  |  |
| 13(a) | Report numbers of individuals at each stage of study | 10 |
| 13(b) | Give reasons for non-participation at each stage | NA |
| 13(c) | Consider use of a flow diagram | 10 |
| 14(a) | Present characteristics of study participants | 12 |
| 14(b) | Indicate number of participants with missing data | Results, first paragraph |
| 15 | Report numbers of outcome events or summary measures | 13 |
| 16(a) | Provide adjusted and unadjusted estimates and precision | 13 |
| 16(b) | Report category boundaries when continuous variables categorized | 12 |
| 16(c) | Translate relative risk estimates into absolute risk where relevant | NA |
| 17 | Report other analyses performed | Results, “Sensitivity analyses”; Supplementary Tables S3–S5 |
| **Discussion** |  |  |
| 18 | Summarise key results with reference to study objectives | 15 |
| 19 | Discuss limitations, sources of potential bias, and imprecision | 18 |
| 20 | Provide a cautious overall interpretation of results | 15 |
| 21 | Discuss generalisability of results | 18 |
| **Other Information** |  |  |
| 22 | Report funding sources and role of funders | 20 |
| 23 | Data availability statement | 19 |
| 24 | Conflict of interest statement | 20 |
| 25 | AI assistance disclosure | 20 |

